# Managing Cyanotic Neonates with Tetralogy of Fallot – A National Perspective on Surgical Approaches?

**DOI:** 10.1101/2025.08.12.25333545

**Authors:** Samuel M Hoenig, David Bruckman, Karl F Welke, Justin Robinson, Rashed Mahboubi, Belinda Udeh, Jarod Dalton, Md M Hossain, Tara Karamlou

## Abstract

**Background:** Management of cyanotic neonates with Tetralogy of Fallot (ToF) remains an important clinical challenge without Level 1 evidence. The Healthcare Cost and Utilization Project Kids Inpatient Dataset (HCUP-KID) is underutilized for evaluating and comparing a national sample of neonates undergoing primary repair, ductal stents, and systemic to pulmonary shunts over time.

**Methods:** Using HCUP-KID, a four-stage algorithm was designed to capture records for neonates with ToF who underwent primary repair, ductal stents, and systemic-to-pulmonary shunts in HCUP-KID years 2016, 2019, and 2022. Hospitalization estimates (n) and percent reported (SE, standard error) reflect weighted results based on survey design. Resource utilization was represented by median hospital length of stay (days) and inflation-adjusted cost in 2023 United States Dollars (USD).

**Results:** An estimated 159.5 discharges nationally were identified for primary repair, 145.3 for ductal stents, and 407.4 discharges undergoing surgical systemic to pulmonary shunt. An estimated 256.3 discharges nationally were identified for infants undergoing definitive repairs following ductal stents. National discharge estimates indicate that ductal stents increased in utilization (linear trend p=0.0011) while surgical shunts decreased (trend p=0.0012). There was an observed decrease in primary repair (trend p = 0.12).

Primary repair had the greatest resource utilization (median LOS 46.9 days and adjusted cost $342,504 2023-USD), followed by surgical shunts (35.1 days, $208,358 2023-USD), ductal stents (21.1 days, $132,259 2023-USD), and definitive repair following ductal stents (7.4 days, $82,788 2023-USD). Between 2016 and 2022, there was a significant increase in the median cost for stent (difference, $77,252 2023-USD, p=0.035) and shunt (difference, $81,111 2023-USD, p=0.043) palliations and an observed decrease in primary repair cost (difference, $76,337, p=0.44).

**Conclusions:** This investigation demonstrates an increased utilization of ductal stents for neonatal ToF across centers nationally. Despite this increase, observed changes in cost reflect a complex paradigm shift that may reflect patient/center-specific decision-making.

## Introduction

Management of cyanotic neonates with Tetralogy of Fallot (ToF) remains an important clinical challenge without clear guidelines or consensus.^1^ Current strategies include neonatal primary repair and staged approaches with either a ductal stent or Blalock Taussig Thomas Shunt (BTTS). To many, the staged approaches are advantageous as they mitigate the immediate cyanotic burden and delay complete repair to the standard infantile window. However, these short-term benefits may not supersede the mid-term ramifications of an unstable interstage period and the cumulative burden of mandatory reintervention or reoperation. Indeed, both ductal stents (DS) and BTTS may require many reinterventions, with the BTTS requiring more reinterventions for cyanosis.^2^ To understand the impact of strategy preference, comparisons of these three strategies have evaluated single-center results,^3^ multicenter clinical data,^4–6^ administrative datasets,^7–11^ and public reporting registries.^12^ Still, the scenario is convoluted, and an assessment of practice patterns and outcomes is needed.

With the increased popularity and integration of DS, this paradigm is evolving. Lemley and colleagues,^13^ in an assessment of the Pediatric Health Information System (PHIS), demonstrated that DS are increasing in popularity over BTTS. Moreover, the COMPASS trial^14^ is currently enrolling cyanotic neonates with ductal-dependent pulmonary flow to assess outcomes between DS and BTTS, but enrollment is stagnating due to lack of equipoise among participating centers. Despite the clinical importance and impact of these two efforts, neither is specific to ToF physiology, and they do not ask the fundamental question: should cyanotic neonates with ToF undergo a primary repair or a staged approach? Further assessment of this paradigm must recognize how such choices impact the clinical and social complexities faced by these patients and their families. As DS become more popular, what are the social factors important for the management of ToF? How should proximity to centers equipped to perform or intervene upon DS impact strategy preference? Finally, is it favorable to choose DS over Primary Repair for symptomatic neonates, recognizing that this increases the healthcare burden on the family responsible for interstage monitoring? To this end, a prospective randomized controlled trial is highly unlikely to be successful, given that this population is small and would likely be underpowered concerning diversity of socioeconomic and racial/ethnic position.

To approach these challenges, our team has worked to establish a decision-analytic framework to study this paradigm from a healthcare economic perspective. Our first study utilized inputs from published studies and found that at a 2-year horizon, from a single-patient clinical perspective, a Primary Repair was preferred insofar as healthcare cost and quality-adjusted life years (QALYs) gained.^15^ Further sensitivity analysis found healthcare costs to be a major factor impacting incremental cost-effectiveness ratios. In the present study, we use the Healthcare Cost and Utilization Project Kids Inpatient Database (HCUP-KID) to define a more complete national sample and understand how the results could inform future iterations of our models.

Through this work, we report national inpatient discharge estimates of cyanotic neonates with ToF requiring surgery for HCUP-KID between the years 2016, 2019, and 2022. We also explore the implications of a paradigm shift where trends in strategy implementation may be associated with changes in resource utilization.

## Methods

A retrospective cohort study was designed to investigate the HCUP-KID. The use of this limited dataset was approved by the Cleveland Clinic Institutional Review Board (IRB# 15-139).

### Data Source and Study Design

HCUP-KID is the largest publicly available pediatric database in the United States derived from discharge billing abstracts, including 48 states and the District of Columbia. It samples 4,000 US hospitals prioritizing “complicated” newborns and other pediatric discharges ages 0 to 20 years at admission. Unweighted, each year contains approximately 3 million pediatric discharges and weighted over 6 million hospitalizations. It is designed to sample 10% of normal newborns and 80% of “other pediatric discharges”. The large sample size enables analysis of “rare conditions (e.g. congenital anomalies) as well as, uncommon treatments (e.g. cardiac surgery).”^16^ This dataset also features many procedural elements (ICD-10-PCS and procedure day since admission) and nonclinical data elements, including payer status and family residence, to enable an assessment of non-clinical patient factors. HCUP-KID was available on a triennial basis, and datasets from years 2016, 2019, and 2022 were utilized to assess the present paradigm.

A four-stage algorithm was designed to capture ToF patients as neonates undergoing primary repair, DS, or SS and ToF infants undergoing complete repair following palliation. **Stage 1** captured patients using ICD-10 diagnosis codes with either a primary ToF diagnosis or associated diagnosis codes (**Figure 1, Supplementary Table 1**). **Stage 2** differentiated neonates from infants using the HCUP-KID neonate identifier. **Stage 3** identified surgical ToF neonates and infants based on the combination and order of procedure codes using procedure day (**Supplementary Table 1**). Initially, the study was designed in 3 stages, yet due to the nature of this limited dataset and data use agreement, it was not feasible to validate the specificity of our algorithm for capturing these patients. Therefore, **Stage 4** was developed.

**Figure 1.**
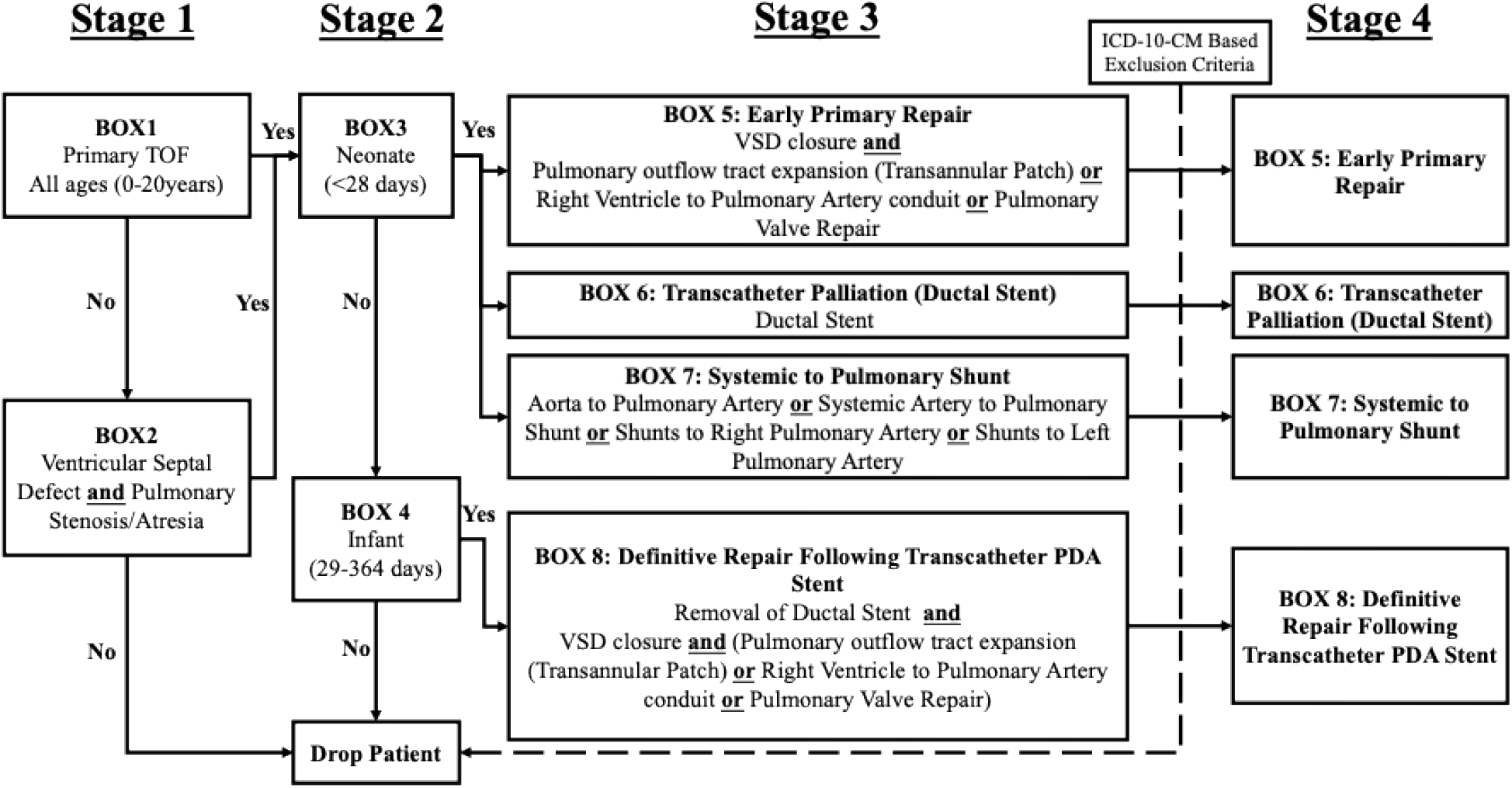
Schematic of the HCUP-KID algorithm designed to define ToF patients undergoing neonatal interventions. A four-stage algorithm was utilized to define neonate tetralogy of Fallot undergoing surgery. Boxes correspond to HCUP-KID specific codes, ICD-10-PCS, and ICD-10-CM codes. Stage 1) Captured discharges for patients with a ToF or comparable diagnosis. Stage 2) Differentiated Neonates from infants. Stage 3) Defined interventional strategies by expected ICD-10 procedural codes for neonates and infants with prior ductal stent palliation. Stage 4) Served as a retroactive validation strategy to improve the specificity of each cohort. *ToF, Tetralogy of Fallot; VSD, Ventricular Septal Defect*.

Using the cohorts defined in Stage 3, reports with counts of all ICD-10 diagnosis codes in each cohort were created. This identified several diagnosis codes that were not consistent with a diagnosis of surgical repair for cyanotic neonates, as adjudicated by three of the investigators (*Investigator Initials: SMH, DB, TK*), with Tetralogy of Fallot (**Supplemental Table 1**). Therefore, to improve the specificity of this algorithm, these diagnosis codes were implemented as final exclusion criteria in Stage 4.

### Study Measures

Descriptive elements, including low birth weight, preterm birth, genetic anomalies, cardiovascular anomalies were identified using ICD-10 codes. Patient family residence was characterized as large (>/= 1 million residents/county) or small (50,000-999,999 residents/county) metropolitan residences, as well as small micropolitan or non-core residences (<50,000 residents/county) using HCUP-KID metrics patient family residence classification Supplementary Table 2). Location of hospital and payer status were also defined using HCUP-KID specific elements (Supplementary Table 2). Outcomes were death, extracorporeal membrane oxygenation, prolonged ventilation (>96 hours), and hospital length of stay (LoS) (Supplementary Table 3). We used the procedure day from admission, provided in HCUP data, to determine when the repair or palliation was performed. Cost estimates (median [Q1, Q3]) were derived from total charges, HCUP cost-to-charge ratio tables and were adjusted to 2023 inflation estimates using the mid-year Consumer Price Index for Medical care.^17^

### Statistical Analysis

To exceed reportable thresholds per HCUP’s data use agreements (N ≥11), all three datasets were combined and weighted appropriately. Hospitalization estimates N, weighted and percent (SE, standard error) reported in tables reflect weighted results based on survey design. Rao-Scott Likelihood Ratio Chi-Square Tests were performed to determine statistical significance between groups. Comparison of costs and LoS across HCUP-KID years used sampling weight-adjusted regression of log-transformed outcomes. Statistical significance of p<0.05 (two-tailed) was used. Pairwise comparisons were adjusted using Tukey method for multiple comparisons and adjusted significance was reported. To test overall changes in surgical strategies, logistic regressions were used to test for overall differences (Wald test) in odds ratios, followed by orthogonal polynomial contrasts for linear trends across years. Sampling weighted adjusted results are presented unless noted.

## Results

We initially identified an estimated 2,196, 2,216, and 2,267 discharges in 2016, 2019 and 2022 respectively, of neonates with a qualifying ToF diagnosis (Box 3, Figure 1). These records were further identified as national inpatient discharge estimates of cyanotic neonates requiring surgery. We captured a nationally weighted estimated 159.5 discharges, patients undergoing Primary Repair (Box 5, Figure 1), 145.3 discharges with DS (Box 6, Figure 1), and 407.4 discharges undergoing a SS (Box 7, Figure 1). 256.3 discharges were identified as having definitive repairs following DS (Box 8, Figure 1; **Table 1**). Patients undergoing definitive repair following a SS palliation could not be reliably identified and were therefore excluded.

**Table 1.** Characteristics of patients captured in HCUP-KID 2016, 2019, and 2022 cross sections combined. Characteristics of Patients undergoing surgery for neonatal ToF.

| Table 1. Characteristics of Patients undergoing surgery for neonatal ToF. |  |  |  |  |
| --- | --- | --- | --- | --- |
|  | Primary Repair | Ductal Stent | Definitive Repair post Ductal Stent | Surgical Shunt |
| Total | N=116<br>(Est. visits = 159.5) | N=107<br>(Est. visits = 145.3) | N=186<br>(Est. visits =256.3) | N=296<br>(Est. visits = 407.4) |
| Patient Clinical Characteristics., % ± se |  |  |  |  |
| Low Birth Weight (<2.5 kg) | 14.4 ± 3.4 | 18.9 ± 4.1 | <6% | 13.7 ± 1.8 |
| Preterm birth (<37 weeks) | 16.3 ± 3.4 | 22.5 ± 4.2 | 5.9 ± 1.7 | 17.1 ± 2.2 |
| Genetic Anomaly | 22.4 ± 4.0 | 15.0 ± 3.8 | 17.3 ± 2.9 | 15.7 ± 2.2 |
| Pulmonary Valve/Artery Atresia | 42.0 ± 4.8 | 52.1 ± 5.2 | 10.2 ± 2.2 | 43.7 ± 3.3 |
| Right Aortic Arch | 0 | 0 | 0 | 0 |
| Tracheal Airway Abnormalities | 9.4 ± 2.6 | <10% | <6% | <5% |
| Patient Urban/Rural Residence., % ± se |  |  |  |  |
| Large Metro, % ± se | 55.7 ± 5.1 | 47.5 ± 5.8 | 59.1 ± 4.1 | 52.4 ± 3.7 |
| Small Metro, % ± se | 31.6 ± 4.8 | 33.8 ± 5.2 | 33.4 ± 3.7 | 34.5 ± 3.4 |
| Micro/Non-core, % ± se | 12.7 ± 3.1 | 18.7 ± 4.2 | 7.5 ± 1.9 | 13.2 ± 2.1 |
| Location/teaching status of hospital, % ± se |  |  |  |  |
| Urban teaching, % ± se | >90% | 100.0 ± 0.00 | >94% | >95% |
| Payer Status, % ± se |  |  |  |  |
| Private Insurance, % ± se | <45% | 42.2 ± 5.9 | 45.6 ± 3.7 | 42.1 ± 3.1 |
| Medicaid, % ± se | 49.1 ± 5.6 | 44.6 ± 6.4 | 43.0 ± 3.8 | 52.8 ± 3.2 |
| Self-pay or other, % ± se | <10% | 13.2 ± 3.9 | 11.4 ± 2.4 | 5.1 ± 1.4 |
| Data suppression applied per HCUP Data Users Agreement. |  |  |  |  |

Characteristics depicting patients’ clinical complexity and potential social determinants of health were defined using ICD-10 diagnosis and HCUP-KID data elements. Nationally, the proportion of neonates with a low birthweight (<2.5 kg) undergoing primary repair, DS, and surgical shunts (SS) was 14.4±3.4, 18.9±4.1, and 13.7±1.8, respectively. A diagnosis of pulmonary artery/valve atresia was present in 42±4.8% of PR, 52.1±5.2% for DS, and 43.7±3.3% for SS. A diagnosis code consistent with genetic anomalies was present in 22.4±4.8%, 15.0±3.8%, and 15.7±2.2% of primary repair, DS, and SS, respectively. No patients had a diagnosis code of right aortic arch. Additional elements of ToF patients’ clinical complexity included preterm birth (<37 weeks) and tracheal airway anomalies and can be found in **Table 1**. A small proportion of patients hailed from micropolitan or non-core residencies, with 12.7±3.1% for primary repair, 18.7±4.2% for DS, and 13.2±2.1% for SS (**Table 1)**. The vast majority of interventions occurred in urban teaching hospitals. Finally, many patients were served by Medicaid insurance with 49.1±5.6% for primary repair, 44.6±6.4% for DS, and 52.8±3.2% for SS (**Table 1**). Additional characteristics can be found in **Table 1**. Overall, patient characteristics were similar, yet due to survey design, comparative statistics were not feasible.

Several hospital binary outcomes often did not exceed the reportable threshold and required suppressed statistics, yet continuous variables, including length of stay and cost, could be reported. In hospital mortality following primary repair was <10%, for DS <8%, and for SS 5.4± 1.2%. Other outcomes including extracorporeal membrane oxygenation and prolonged ventilation (>96 hours), also did not exceed this threshold for primary repair and DS, and the suppressed estimates can be found in **Table 2**. Concerning median hospital LoS and inflation-adjusted cost of hospitalization at the initial admission, primary repair was the most substantial (46.9 days, $342,504 2023-USD), followed by SS (35.1 days, $208,358 2023-USD), DSs (21.1 days, $132,259 2023-USD), and finally, definitive repair following DS (7.4 days, $82,788 2023-USD) **(Table 2)**. In HCUP-KID, assessment of interstage re-admissions could not be assessed. While binary outcomes were challenging to assess, continuous variables showed that primary repair had the most notable resource utilization.

**Table 2.** Outcomes, hospital length of stay, and resource utilization derived from HCUP-KID. *ECMO, extracorporeal membrane oxygenation*.

|  | Primary Repair | Ductal Stent | Definitive Repair following Ductal Stent | Surgical Shunt |
| --- | --- | --- | --- | --- |
| Total | N=116<br>Est. visits =159.5 | N=107<br>Est. visits = 145.3 | N=186<br>Est. visits =256.3 | N=296<br>Est. visits = 407.4 |
| Binary Outcomes for each Strategy, % ± se |  |  |  |  |
| Died during hospitalization | <10% | <8% | <6% | 5.4 ± 1.2 |
| ECMO | <10% | <8% | <6% | 8.1 ± 1.5 |
| Prolonged Vent >96h | <10% | <8% | <6% | 4.9 ± 1.3 |
| Total Resource Utilization of each strategy, median [Q1, Q3] |  |  |  |  |
| Length of stay | 46.9<br>[22.9, 88.8] | 21.1<br>[13.4, 47.5] | 7.4<br>[5.0, 13.8] | 35.1<br>[20.5, 58.9] |
| Total cost adj to 2023 USD | 342504<br>[147799, 666220] | 132259<br>[82524, 278881] | 82788<br>[60581, 131322] | 208358<br>[132801, 361213] |
| Statistics are weighted. Data suppression applied per HCUP Data Users Agreement. |  |  |  |  |
| SAS Survey Procedures used for all analyses. |  |  |  |  |

Within this timeframe, there were substantial changes in strategy utilization. For palliative approaches, the estimated number of ductal stent discharges increased (n= 26.7 in 2016 to n = 77.1 in 2022), and the estimated number of SS decreased (n = 184.9 in 2016 to n = 103.5 in 2022). Moreover, neonates admitted for ToF were 2.86 times more likely to have had a ductal stent in 2022 versus 2016 (OR=2.86, 95% confidence interval (1.52, 5.39). Although there was not a significant difference in likelihood of a ductal stent in 2019 versus 2016 (OR=0.55 (0.78, 3.09)), a linear trend was observed (p=0.0011) over the three cross-sectional years. The estimated number of SS discharges decreased from 184.9 in 2016, 119.0 in 2019, to 103.5 in 2022. Neonates in 2022 with ToF were 38% times likely (OR=0.62 (0.44, 0.86) in 2022 than 2019 and 48% as likely (OR=0.52 (0.35, 0.77) to receive SS compared to 2016, and (n= 26.7 in 2016 to n = 77.1 in 2022; linear trend p=0.0012) (**Figure 2**). Sampling weight-adjusted number of discharges for neonates undergoing early primary repair changed from 59.5 in 2016 to 61.9 in 2019 and to 38.1 in 2022. But neonates with ToF had statistically insignificant odds of undergoing primary repair in 2022 versus 2016 (OR=0.61 (0.33, 1.14)) or in 2019 versus 2016 (OR=1.03 (0.59, 1.82), linear trend p=0.12). Though only observational, between 2019 and 2022, neonates were less likely to undergo definitive repair (OR=0.59 (0.33, 1.06), p>0.05) (**Figure 2)**. Such results suggest that DS may be gaining in popularity over surgical options.

**Figure 2.**
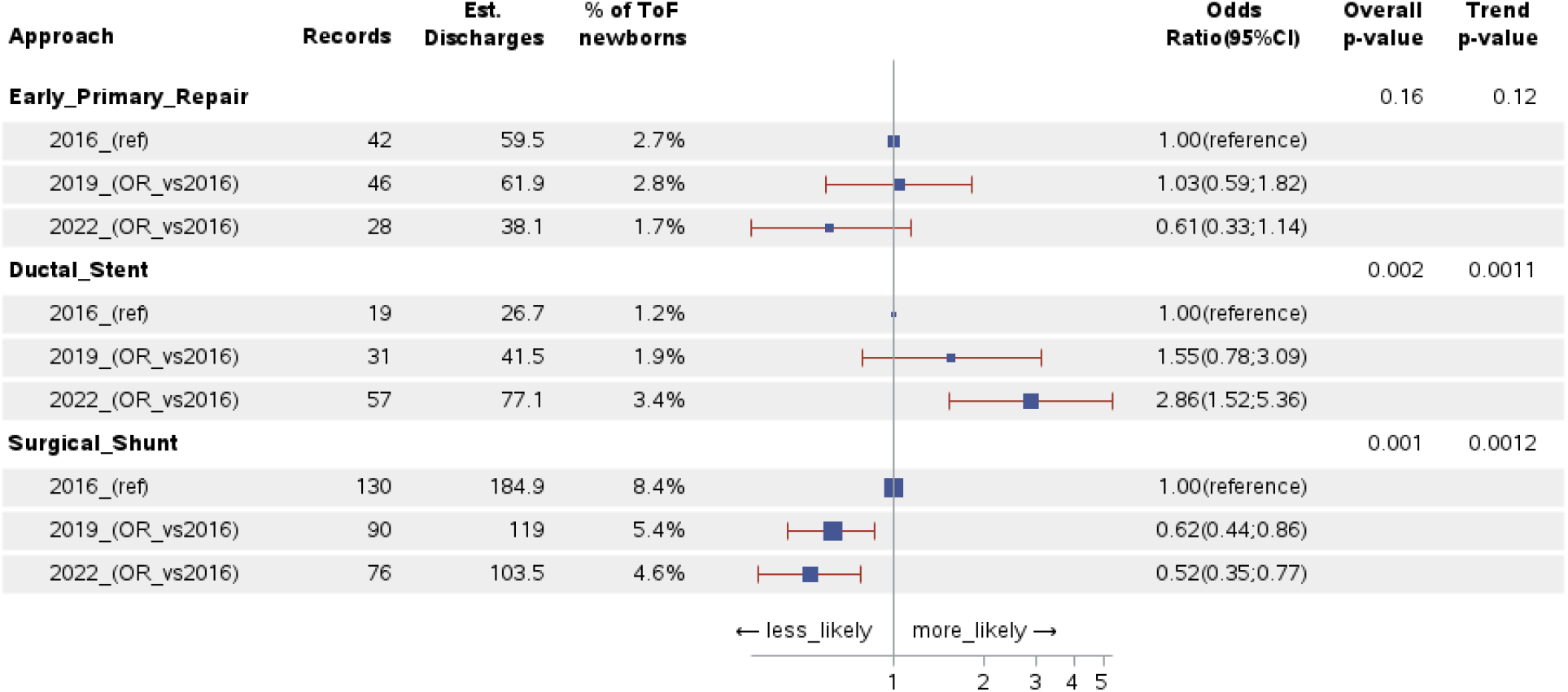
Odds of neonatal tetralogy of Fallot intervention occurring in HCUP-KID since 2016. Forest plot depicting odds of neonatal tetralogy of Fallot intervention occurring in HCUP-KID. *Primary Repair:* Between 2019 and 2016, there was no observed change in the odds of primary repair (OR=1.03 (0.59, 1.82). Yet between 2019 and 2022, there was an observed decrease in the odds of primary repair (OR=0.59 (0.33, 1.06), p>0.05). The total decrease in primary repair utilization from 2016 to 2022 was not statistically significant (OR=0.61 (0.33, 1.14), linear trend p=0.12). *Ductal Stent*: From 2016 to 2022, neonates were more likely to have had a ductal stent (OR=2.86, 95% confidence interval (1.52, 5.39) with a linear trend (p=0.0011). *Surgical Shunt:* Compared to 2016, neonates in 2019 were 38% times as likely to receive a surgical shunt (OR=0.62 (0.44, 0.86), and in 2022, 48% as likely (OR=0.52 (0.35, 0.77) witha linear trend (p=0.0012). These results suggest that since 2016, ductal stents have gained in popularity, and surgical shunt utilization has decreased. The observed decrease in primary repair may also be clinically significant.

With the changes in strategy across the three cross-sections, there were observed changes in median hospital LoS and inflation-adjusted cost. From 2016 to 2022, median LoS for primary repair decreased by 8.6 days (overall p=0.67). Median total costs decreased by 21% ($76,337 2023 USD). This difference was primarily between 2019 and 2022, but overall, assessing a linear trend, these differences across the three years were observational (p=0.44, overall). For DS, median total costs increased significantly (difference in medians, $77,252 2023-USD, overall p=0.043, pairwise p=0.035), paralleled by an observed 7.4 day increase in median LoS (overall p=0.46). The total cost for SS also increased significantly (difference in medians, $81,111 2023-USD, overall p=0.0446, pairwise p=0.043), with an observed 12.5 day increase in hospital LoS (marginally significant at p=0.058). Between 2016 and 2022, there was no significant change in costs for definitive repair following DS (difference in medians, $11,584 2023-USD, overall p=0.65 and 0.4 days, overall p=0.80) (**Table 3**). Further inquiry into ductal stent and SS hospitalizations found that these palliative strategies were consistently performed early in the hospitalization and did not significantly change between the three cross sections (**Table 4**). While the change in resource utilization for primary repair is not statistically significant, there has been a significant increase in resource utilization for palliative strategies.

**Table 3.** Model-derived median values depicting trends in hospital length of stay and resource utilization as noted by cost between strategies. Statistical significance derived by Turkey adjusted p-values indicates statistical significance between cross sections noted by # and ##.

| Year | 2016 | 2019 | 2022 |
| --- | --- | --- | --- |
| Median Hospital Length of Stay (Days) |  |  |  |
| Early Primary Repair | 54.3 | 48.3 | 45.7 |
| Ductal Stent | 21.7 | 25.6 | 29.1 |
| Definitive Repair following Ductal stent | 11.4 | 10.4 | 11.1 |
| Surgical Shunt | 34.2 | 37.6 | 46.7 |
| Median Cost (2023 USD) |  |  |  |
| Early Primary Repair | \$362,869 | \$360,723 | \$286,532 |
| Ductal Stent | <u>\$97,051</u><sup>#</sup> | \$163,158 | <u>\$174,303</u><sup>#</sup> |
| Definitive Repair following Ductal stent | \$91,394 | \$96,162 | \$102,978 |
| Surgical Shunt | <u>\$208,414</u><sup>##</sup> | \$213,481 | <u>\$289,525</u><sup>##</sup> |
| #: $p = 0.0347$ for the 2016 vs 2022 comparison<br>##: $p = 0.0429$ for the 2016 vs 2022 comparison | | | |

**Table 4.** Trends in the day of neonatal interventional strategy between the three cross-sectional time points.

| Strategy | Year | N | Estimated Discharges (N weighted) | Day Intervention Occurred |  |  |  |  |
| --- | --- | --- | --- | --- | --- | --- | --- | --- |
|  |  |  |  | Median | Quartile 1 | Quartile 3 | Min | Max |
| Early Primary Repair | 2016 | 42 | 59.5 | 4.8 | 3.09 | 8.25 | 1 | 13 |
|  | 2019 | 46 | 61.9 | 4.2 | 2.20 | 8.60 | 1 | 13 |
|  | 2022 | 28 | 38.1 | 5.7 | 3.49 | 8.73 | 1 | 20 |
| Ductal Stents | 2016 | 19 | 26.7 | 3.1 | 1.42 | 4.66 | 1 | 15 |
|  | 2019 | 31 | 41.5 | 3.6 | 1.65 | 6.04 | 1 | 19 |
|  | 2022 | 57 | 77.1 | 2.9 | 1.15 | 5.74 | 1 | 18 |
| Surgical Shunts | 2016 | 130 | 184.9 | 2.8 | 1.00 | 5.27 | 1 | 15 |
|  | 2019 | 90 | 119 | 3.5 | 1.80 | 6.55 | 1 | 18 |
|  | 2022 | 76 | 103.5 | 3.9 | 1.43 | 7.66 | 1 | 20 |

## Discussion

The present study builds on our prior work using decision analysis to determine the optimal treatment for the cyanotic neonate with ToF. Here, we have investigated source data rather than published probabilities and developed algorithms for cohort identification that can be utilized more broadly to investigate challenges and changes within CHS. We show that 1) through this framework, HCUP-KID with the ICD-10 can be used to define a national dataset of cyanotic ToF neonates requiring surgery, and also capture infants undergoing complete repair following DS. 2) From 2016 to 2022, DS significantly increased in utilization while surgical palliation decreased. We also observed a trend toward a decrease in primary repair. 3) With this change, there was an observed reduction in cost for primary repair and significant increases in cost of palliative strategies. These trends may reflect risk diversion where higher-risk ToF neonates are treated with DS rather than primary repair and therefore accruing a greater cost for care. Alternatively, it could be simply that many more centers, regardless of surgical volume, are opting for DS in cyanotic neonates, and triaging higher-risk neonates (those with hypoplastic pulmonary arteries or other comorbidities/syndromes) to higher volume centers for primary repair, where costs may be lower.

### Capturing a National Sample

We integrated ICD-10 diagnosis and procedure codes as well as the order of events based on procedure day (reported in 98% of ToF study records) or column order to capture a national sample of ToF neonates requiring intervention. Similar ICD strategies have been employed in other datasets (PHIS and HCUP-National Inpatient Sample) and clinical registries (STS-CHSD).^7,10,11,18,19^ However, assessment of neonatal palliative strategies has not been attempted in HCUP-KID. Our novel choice to combine ICD-10 codes that reflect ligation or removal of a patent ductus arteriosus with codes for definitive repair also allowed us to differentiate those undergoing definitive repair post-ductal stent placement from infants undergoing an index definitive repair. Therefore, across this dataset, major steps of the ductal stent pathway could be captured without linkage codes that could identify serial admissions. Concerning prior investigations, Al Habib and colleagues^12^ across 6 years of the STS-CHSD (2002-2007) captured 154 neonates undergoing primary repair for ToF and 178 SS. Savla and colleagues,^10,11^ using 12 years of PHIS (2004-2015) captured 1,032 neonates undergoing primary repair and 1,331 patients undergoing surgical palliation. At the time of these studies, DS for ToF were novel and not defined in the ICD-9. We captured a comparable or greater number of surgical patients in each given timeframe. The present study builds upon prior administrative investigations into the neonatal ToF paradigm, with results consistent with prior data and the integration of the ductal stent pathway.

### Paradigm Shift in Utilization and Cost

This analysis highlights a paradigm shift. From 2016-2022, patients receiving DS increased, which was reciprocated by decreases in SS. These results are consistent with Lemleys^13^ experience with PHIS. However, our results also suggest that between 2016 and 2019 DS utilization may not have had an impact on primary repair preference. Then, between 2019 and 2022, DS may have gained popularity over primary repair. While this trend was not statistically significant, it is clinically relevant and consequential.

Initially, we hypothesized that as the ductal stent learning curve was superseded, the utilization of hospital resources would decrease and be reflected by a decreased total cost and hospital LoS. Surprisingly, as DS gained popularity, we observed an increased cost for both palliative strategies, and similarly, costs of primary repair decreased, coincident with decreased primary repair utilization. Such trends could not be explained by differences in rudimentary patient characteristics defined using ICD-10 diagnoses or a longer clinical preamble to the index procedure. Moreover, as HCUP-KID is derived from claims data and costs from a composite charge value, further investigations into the drivers of cost were not feasible. We speculate that as DS have been broadly implemented, the armamentarium of strategies offered has expanded. As a result, centers may be choosing higher-risk neonates for a ductal stent palliation, incurring a greater cost. Moreover, high-volume centers may also be developing center-specific clinical and personalized criteria, potentially reflecting a salutary benefit for patients selected to undergo primary repair. This may explain why the observed cost of primary repair has decreased.

Furthermore, as DS are adopted into small center practice, they may be utilized over primary repair, which is then selectively shifted to high-volume centers. These low-volume centers may also have less experience with DS and incur a greater cost of care. This broad implementation and resulting sub-optimal results are well understood as the *Trough of Disillusionment* in the Gartner hype cycle.^20^ Further investigation into the ductal stent learning curve is necessary to improve resource utilization and to understand the optimal strategy for each patient.

Despite a potential shift to personalized care, these changes raise concerns regarding the high financial burden that palliative strategies place on patients and their families. DS are noted for a high incidence of reintervention. Up to 84% require a subsequent balloon angioplasty, and ~16% may require an additional stent.^2^ This may require additional time off work, travel, and childcare costs for families. Moreover, Welke and colleagues^21^ found that the median travel distance from a patient’s home address to a CHS center was 38.5 miles (IQR 14.7 – 102 miles), with 25% traveling over 100 miles, and 53% choosing a hospital outside their region. This context is important, as the initial ductal stent and subsequent interventions may not be offered at local or low-volume centers. In such instances, a definitive repair would enable a child to be discharged home, to rural, distant, or disenfranchised residences, without the need for additional immediate reinterventions. Indeed, these social factors have contributed to the decision-making process at our centers, and future studies weighing this choice are necessary to understand the social perspective of this complex paradigm shift and to deliver personalized, high-value care.

### High Financial Burden of Neonatal Primary Repair

In our prior cost-effectiveness analysis, we used inflation-adjusted costs, derived from clinical data linked with PHIS.^4^ At baseline, we found that primary repair was more cost-effective than either transcatheter stents or SS. In the present study, the median hospitalization cost for primary repair was substantially larger than the estimates incorporated into our models. The primary repair cost was also greater than the cost of individual palliative strategies and the sum of DS and subsequent definitive repair. These results oppose prior PHIS data,^9^ which demonstrated a significantly greater linked cost for the entire palliative pathway compared to a single-stage repair for cyanotic ToF neonates. The present study also varies from O’Byrne and the Congenital Cardiac Research Collaborative,^4^ who merged 18 months of clinical data in 6 high-volume centers with PHIS. After inflation-adjusting from 2017 to 2023 USD,^17^ the accrued 18-month cost of the primary repair pathway from O’Byrne’s study ($221,131 2023-USD) is lower than the median cost from the primary repair hospitalization found in HCUP-KID. Such differences are unsurprising given that HCUP-KID represents large and small volume programs, and variation in resource utilization based on volume in CHS is well-documented.^22^ High-volume centers of excellence show lower resource utilization for complex care and may not be reflective of national practice. Future iterations of our models and other assessments of the cost of CHS require cost estimates from similarly large and representative samples to fully appreciate the financial implications in value-based care.

### Lessons Learned from HCUP-KID

The present study is the second assessing the cyanotic neonatal ToF paradigm in HCUP-KID^23^ and the first utilizing ICD-10 codes to define the three major palliative ToF strategies. This source data has proven valuable in understanding trends in cost and LoS, yet underscores important challenges to be considered for future assessments.

### Assessing Binary Outcomes

Quantification of binary outcomes in this study was challenging. This may be because the incidence of cyanotic neonatal ToF in three years was low, and because the majority of morbidity and mortality for these patients occurred after discharge.^6^ Moreover, the management of ToF has greatly improved in recent decades, with an estimated survival approaching 100% for standard-risk infants^24^ and >90% for cyanotic neonates at centers of excellence.^6^ With the present outcomes, other measures of morbidity have a low incidence and may not exceed the reportable threshold for a small sample. For instance, Ghimire and colleagues^23^ compared cyanotic ToF neonates to standard-risk ToF infants in HCUP-KID 2003-2012. They captured a large cohort using a now-discontinued code for complete ToF repair (ICD-9 35.81). In this antiquated cohort, binary outcomes such as mortality, extracorporeal membrane oxygenation, and respiratory failure were reported in incidences < 10% and only exceeded the reportable threshold due to the high sensitivity of the prior code. For a sample in the ICD-10, comparing multiple neonatal ToF interventions, a comparison of such outcomes was not feasible.

Moreover, HCUP-KID truncates patients’ core files at 40 diagnosis codes and 25 procedure codes.^25^ Additional codes reflecting morbidity for complex patients may further be omitted due to the complexity of these patients and the limitations of the registry design.

### Characterizing surgical strategies in HCUP-KID

Due to the file design, deterministic identification of serial hospitalizations was not feasible. As such, we characterized infants receiving definitive repairs who had received a prior ductal stent using procedure codes that would reflect a removal of the stent. We attribute this success to the ICD-10 transition, where codes for ductal stent placement/removal were clearly articulated. However, attempts to define definitive repair for patients following SS were unsuccessful.

Three strategies were attempted to capture infant definitive repairs following SS. First, emulating the capture of ductal stent infants, procedural codes that might indicate the removal of a SS from the pulmonary arteries were applied with codes for a definitive repair (Supplementary Table 4, Supplementary Figure 1). While unsuccessful, presumed shunt take-down codes were substituted for diagnosis codes that might indicate prior cardiac surgery or repair of the sternum or chest wall (Supplementary Table 4). Both strategies resulted in the capture of patients either disproportional to surgically palliated ToF patients or below the reportable threshold (N<11). Out of options, our team assessed reports of ICD-10 procedure and diagnosis codes for neonates and infants captured in stage 2 of the algorithm. We were surprised to find a large incidence of codes describing a thymectomy. We rationalized that the absence of a Thymectomy code could indicate a prior median sternotomy for SS in this population (Supplementary Figure 2, Supplementary Table 4). Before the application of this theory, we assessed the incidence of Thymectomy reporting for ToF neonates previously captured in stages 1-3. These codes were only identifiable in a fraction of patients and present for some with a DiGeorge diagnosis. These preliminary findings indicated that this strategy had low sensitivity and specificity, indicating that a thymectomy code cannot be used to indicate a prior sternotomy. Our team has three main explanations for these challenges, detailed in the **supplemental discussion**. With these challenges, our team further investigated the HCUP-National Readmission Dataset, which possessed linkage codes and showed promise for identifying previously shunted patients. Deterministic characterization of procedure order within this dataset presented new and unique challenges precluding a full assessment. These are detailed in the **supplemental discussion**.

While partially successful, our efforts to define definitive repair patients based on these strategies hold promise for future investigations. HCUP is derived from State Inpatient Databases; therefore, the representation of complex care paradigms that cross state lines is limited. This proved to be a significant challenge in our investigations into the national readmission dataset. Also, readmissions and reinterventions that crossed state lines could not be identified. Our novel strategy leveraging procedure day offers the opportunity to capture patients with a prior ductal stent and utilize HCUP for assessing national practice patterns for surgical pathways in CHS.

### Dataset validation

Validation of captured patients with clinical data is not routinely performed in HCUP studies, including prior assessments for this paradigm.^7,23^ Even so, we sought to understand and improve the specificity of our cohort and retroactively developed stage 4. In this process, we were surprised to find that some patients carried diagnosis codes inconsistent with the ToF paradigm. For example, multiple patients had a hypoplastic left heart syndrome diagnosis (Q23.4). This can be explained by the complexity and heterogeneity of each defect; some patients with severe ToF and Double Outlet Right Ventricle (DORV) physiology are managed on a single ventricle pathway. Initially, their charts may include both diagnoses. This coding error may be further exacerbated in our algorithm as the BTTS is routinely used in the Norwood procedure and DS are essential for hybrid palliation. Allen and colleagues^18^ recently developed a publicly available risk stratification tool (RACHS-2) for CHS using PHIS and STS-CHSD. With similar ICD-10 codes, they partitioned complete ToF repair and SS with excellent specificity.

However, this study did not assess SS specifically in neonatal ToF patients, nor did they highlight the utility of DS in this scenario. These nuances are important for working with specific CHS populations that are not routinely studied on a national scale. For future investigations into HCUP-KID, when external validation cannot be achieved, rigorous internal validation for misdiagnosed patients is essential.

### Additional Limitations

This is a limited dataset, advertised for promise evaluating CHS patients;^16^ however, it was challenging to capture a small population. Additionally, the sampling design, focusing on 80% of “complicated” pediatric discharges, potentially omitted many cyanotic ToF neonates who could have skewed results. Future studies into HCUP-KID may benefit from larger, less specific populations of CHS patients. Additionally, assessing hospital discharges serves as a snapshot of care for these complex patients and is not a complete representation of the paradigm. A longitudinal assessment of resource utilization in this paradigm may not be feasible with administrative data; future studies leveraging analytical models or prospective cohorts^26^ may be necessary to improve value-based care for these patients and their families.

## Conclusions

The present study builds on our prior work defining value-based care for cyanotic neonates requiring surgery for ToF. Utilization of DS is increasing. This is reciprocated by a decrease in SS and primary ToF repairs. This trend may reflect risk diversion, thereby prolonging the ‘hype cycle’ for the ductal stent innovation, further reflected by an increase in the resource utilization for palliative strategies and a potential decrease in the resource utilization for primary repair.

These findings suggest that centers may be choosing individualized clinical benchmarks in their decision-making, and more investigations are necessary to understand this complex paradigm shift.

## Data Availability

Data may be shared upon request to the coresponding author following and agreement from multi-institutional statistical team.

## Acronyms and Abbreviations

ToF: Tetralogy of Fallot
BTTS: Blalock Taussig Thomas Shunt
HCUP: Healthcare Cost and Utilization Project
PHIS: Pediatric Health Information System
KID: Kids Inpatient Dataset
LoS: Length of Stay
USD: United States Dollars
CHS: Congenital Heart Surgery
DS: Ductal Stent
SS: Surgical Shunt

## Acknowledgments

We are immensely grateful to the Center for Populations Health Research, Lerner Research Institute, Cleveland Clinic for their collaboration and funding through this project. We would like to acknowledge the work of the United States Agency for Healthcare Quality and Research for collating and providing access to Healthcare Cost and Utilization Project’s Kids Inpatient Dataset and National Readmission Sample. We would also like to thank the states that contributed to the AHQR in their collation of the HCUP datasets.

